# Predicting the Predictable in the Psychiatric High Risk

**DOI:** 10.1101/2025.04.11.25325553

**Authors:** Eric V. Strobl

## Abstract

Most investigators in precision psychiatry force models to predict clinically meaningful but ultimately predefined outcomes in a high-risk population. We instead advocate for an alternative approach: let the data reveal which symptoms are predictable with high accuracy and then assess whether those predictable symptoms warrant early intervention. We correspondingly introduce the Sparse Canonical Outcome REgression (SCORE) algorithm, which combines items from clinical rating scales into severity scores that maximize predictability across time. Our findings show that this simple shift in perspective significantly boosts prognostic accuracy, uncovering predictable symptom profiles such as social difficulties and stress-paranoia from those at clinical high risk for psychosis, and social passivity from infants at genetic high risk for autism. The predictable scores differ markedly from conventional clinical metrics and offer clinicians memorable, actionable insights even when full diagnostic criteria are unmet.

## 1. Introduction

A population of high-risk patients refers to a set of individuals with an elevated likelihood of developing a mental illness based on a variety of factors such as early symptoms, genetic predispositions, or environmental exposures (Yung and McGorry, 1996). High-risk patients represent an enriched group that justify detailed consideration despite limited clinical resources (McGorry et al., 2006). However, predicting which individuals will develop mental illness within a high-risk sub-population remains a persistent challenge.

Failed predictions delay interventions, worsen patient outcomes, and strain overburdened mental health systems – underscoring the need for effective approaches (Fusar-Poli et al., 2017). As a result, many investigators have resorted to complex machine learning models to forecast the development of mental illnesses (Chekroud et al., 2021). Numerous models have unfortunately achieved only modest accuracy despite incorporating tens or even hundreds of predictors (Chekroud et al., 2024). The problem has in turn caused investigators to explore additional predictors or adopt cutting-edge algorithms, escalating model complexity in a reinforcing cycle. Investigators hope that at least one model will eventually deliver high accuracy – an assumption possibly inspired by machine learning culture, where tasks like object differentiation in images are known to be predictable a priori, so accuracy primarily hinges on sufficient model complexity (Feng et al., 2021).

In medicine, however, some outcomes may simply defy prediction, especially in longer time horizons (Simon et al., 2022). As a result, forcing algorithms to predict a clinically important but ultimately fixed outcome may stubbornly yield unsatisfying results. For instance, accurately predicting a patient’s depression level two years into the future may prove elusive due to random changes in living conditions, social situations, or access to therapy. We therefore believe that algorithms should not only predict the future but also identify what can actually be predicted among a set of multiple outcomes. Clinicians can then interpret post hoc which predictable outcomes merit clinical action. To achieve this, we analyze the problem from first principles and design a tailored method that extracts novel insights from high-quality and purposefully built datasets, rather than reinforcing further model complexity with a sample-hungry algorithm that relies on very large datasets originally collected for unrelated goals. We thus prioritize predicting the predictable through a seemingly simple but meticulously designed approach customized for the demands of precision psychiatry.

We, in particular, advocate for learning predictable *summary scores* that combine many individual items of a clinical rating scale into a few composite metrics, rather than predicting each item. To illustrate, consider an extreme case where a rating scale contains hundreds of individual items that would quickly overwhelm clinicians if we predicted each item separately. In contrast, consolidating hundreds of items into a 2-5 summary scores enhances interpretability. Further, consolidation can increase predictability beyond any single item by leveraging multivariate relationships between items. Learned summary scores thus effectively balance statistical complexity with clinical interpretability.

Existing total scores and sub-scores of most clinical rating scales are unfortunately designed to summarize patients in the here and now rather than to maximize prognostic accuracy (Baer and Blais, 2010). We therefore make the following contributions in this paper:

1. We propose to *non-negatively* weigh the many individual items in a clinical rating scale to create a few summary *severity* scores that are maximally predictable across a time interval (Figure 1).
2. We specifically define predictability as the ability to differentiate between patients in a high-risk group by maximizing correlation, rather than minimizing the absolute error.
3. We correspondingly introduce the Sparse Canonical Outcome REgression (SCORE) algorithm that learns the summary scores from longitudinal data by maximizing the correlation.
4. We find that total scores and clinical sub-scores are sub-optimal for prediction, and predictable summary scores differ substantially from total scores and subscores in both psychosis and autism spectrum disorder (ASD).
5. We also find that summary scores differ in their predictability depending on the time horizon, and some predictable summary scores warrant clinical action.

We therefore conclude that outcome measures should, just like predictors, be subjected to machine learning transformations and clinically interpreted post hoc – rather than kept fixed.

**Figure 1.**
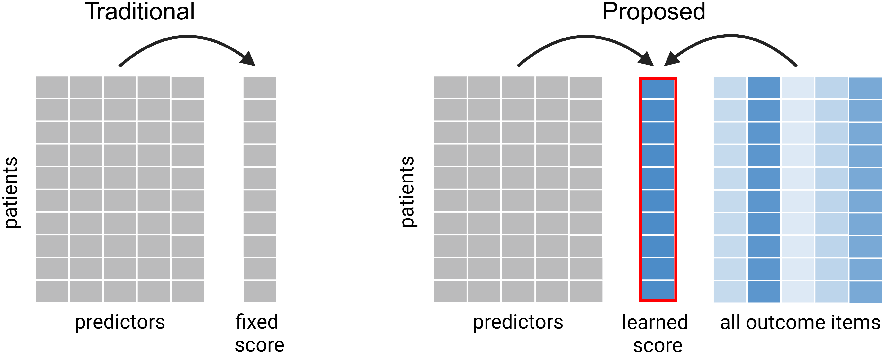
Traditional methods force a machine learning algorithm to predict a fixed outcome (left). We propose to optimally combine all individual items in a clinical rating scale to learn the most predictable summary outcome across time (right). Though not depicted, we further weight the outcome items multiple times to learn all predictable summary scores.

### Generalizable insights

The SCORE algorithm generalizes to numerous complex illnesses beyond its focus on psychiatric high-risk populations. Metabolic disorders and rheumatologic diseases, for example, exhibit diverse, interconnected symptoms that evolve dynamically (McInnes and Schett, 2011). These conditions resist reduction to a single score and instead require multiple individual items – like blood biomarkers, joint inflammation markers, and patient-reported outcomes – to capture their complexity. SCORE integrates these variables into interpretable severity scores that maximize predictability. The algorithm pinpoints forecastable disease aspects instead of forcing models to predict predefined, often elusive, outcomes. For example, SCORE might highlight predictable combinations of insulin resistance and neuropathy in diabetes, while forecasting joint-specific inflammation and pain scores in rheumatoid arthritis. The approach thus shifts the paradigm from forcing predictions to predicting what can actually be anticipated.

## 2. Related Work

Investigators traditionally distinguish patients in high-risk populations by training machine learning models to predict a fixed, clinically meaningful outcome measure. For example, investigators often attempt to predict the conversion to psychosis, a diagnosis defined by the Structured Interview for Psychosis risk Syndromes (SIPS) based primarily on positive symptoms (Cannon et al., 2016). These symptoms, such as hallucinations and delusions, exhibit the highest specificity to psychosis. In contrast, cognitive impairments or negative symptoms, such as blunted emotions, reduced speech, and difficulty with motivation, often overlap with other conditions, reducing their utility as primary conversion criteria (McCutcheon et al., 2023; Mosolov and Yaltonskaya, 2022). Likewise, autism diagnosis in toddlerhood targets social affect and restricted or repetitive behaviors due to their specificity to the disorder (Lord et al., 2012). Existing criteria thus prioritize symptoms unique to each diagnostic category.

Symptoms optimal for diagnosis are not necessarily optimal for other clinical tasks like treatment assignment, subgroup differentiation or – as emphasized in this paper – prognostication. As a result, investigators have developed methods that explicitly learn optimized outcome measures for specific tasks in psychiatry. The Supervised Varimax algorithm, for instance, rotates an orthogonal set of factors to maximally differentiate the causal effects of treatments (Strobl and Kim, 2024; Strobl, 2024). The algorithm has also been adapted to distinguish between predefined populations, such as those with ASD versus healthy controls (Strobl, 2025). However, Supervised Varimax and its variants prioritize differentiating treatment effects or predefined subgroups over developing severity scores that maximally predict individual patient trajectories. As a result, they solve fundamentally different tasks. The algorithms also cannot admit non-negativity constraints essential for learning predictable *severity* scores.

Investigators in the machine learning community have also developed methods to optimally transform outcome measures. For example, the Alternating Conditional Expectations (ACE) algorithm minimizes the mean squared error (MSE) between outcomes and predictors through two alternating regressions (Breiman and Friedman, 1985). One regression predicts outcomes using predictors, while the other predicts predictors using outcomes. ACE maximizes the correlation between predictors and outcome with one outcome variable, but otherwise prioritizes MSE reduction with more than one outcome. Unfortunately, minimizing the MSE biases the algorithm to select stable symptoms that yield precise predictions rather than differentiating patients with worsening symptom severity (Section 3). We thus need to carefully distinguish machine learning accuracy metrics from clinically useful ones.

Another method called Non-negative Canonical Correlation Analysis (NCCA) partially addresses the above issue by maximizing correlation for the first canonical variate, even with multiple outcomes, rather than minimizing the MSE (Sigg et al., 2007). The non-negativity constraint on clinical rating scale items ensures that the weighted metric corresponds to a severity score. However, NCCA also adheres to traditional canonical correlation analysis by enforcing (approximate) uncorrelatedness among canonical variates (Mackey, 2008). Enforcing uncorrelatedness may prevent the algorithm from capturing clinically relevant symptom profiles. For example, social withdrawal and anxiety symptoms correlate, so NCCA may discover a severity score involving negative symptoms in one canonical variate but underrepresent anxiety symptoms in later variates, even though anxiety is highly treatable. Enforcing uncorrelatedness therefore risks missing symptoms that are clinically useful to identify.

Investigators have finally applied unsupervised methods, such as hierarchical symptom clustering and non-negative matrix factorization (NMF), to learn symptom severity scores (Lee and Seung, 2000; Chekroud et al., 2017). However, these approaches may not prioritize *predictable* summary scores, especially those that explain smaller proportions of the variability in the individual items. Supervision is therefore critical for discovering all predictable summary scores.

In summary, existing methods fail to comprehensively learn symptom severity scores that maximize predictability. The algorithms leave clinical utility incomplete by either prioritizing specific symptoms, overlooking individual patient differentiation, forcing uncorrelatedness among canonical variates, or failing to account for independent variables.

## 3. Predictability as Distinguishing Patients

We must first carefully define predictability to suit our clinical objective. In a high-risk patient cohort, we aim to distinguish individuals whose symptom severity will worsen from those whose will not. We prioritize identifying differences among patients within this group over predicting symptom severity across the entire high-risk population.

Standard regression metrics like the MSE, mean absolute deviation and Huber loss focus on minimizing the deviation between predicted and actual values (Hastie et al., 2009). The MSE, for example, penalizes larger errors quadratically, emphasizing overall accuracy but potentially favoring models that predict stable outcomes with low error – even if they fail to capture relative differences among individuals. The mean absolute error penalizes errors linearly but still prioritizes absolute accuracy. Similarly, the Huber loss blends quadratic and linear penalties but focuses on error reduction. These metrics thus cause algorithms to converge on solutions that yield precise predictions but lack the power to separate individuals effectively within the high-risk group.

In contrast, the Pearson correlation coefficient measures the strength and direction of the linear relationship between predicted and actual values. Correlation thus reflects how well predictions rank individuals linearly, rather than focusing on absolute error. As a result, correlation is better suited for *distinguishing* patients within a cohort whose outcomes diverge. For example, minimizing the MSE might lead to a model that predicts a narrow range of stable outcomes with low variance and error, while maximizing the correlation highlights a model’s ability to order patients by their true outcomes, even if absolute predictions are less precise (Figure 2 (a) and (b)).

**Figure 2.**
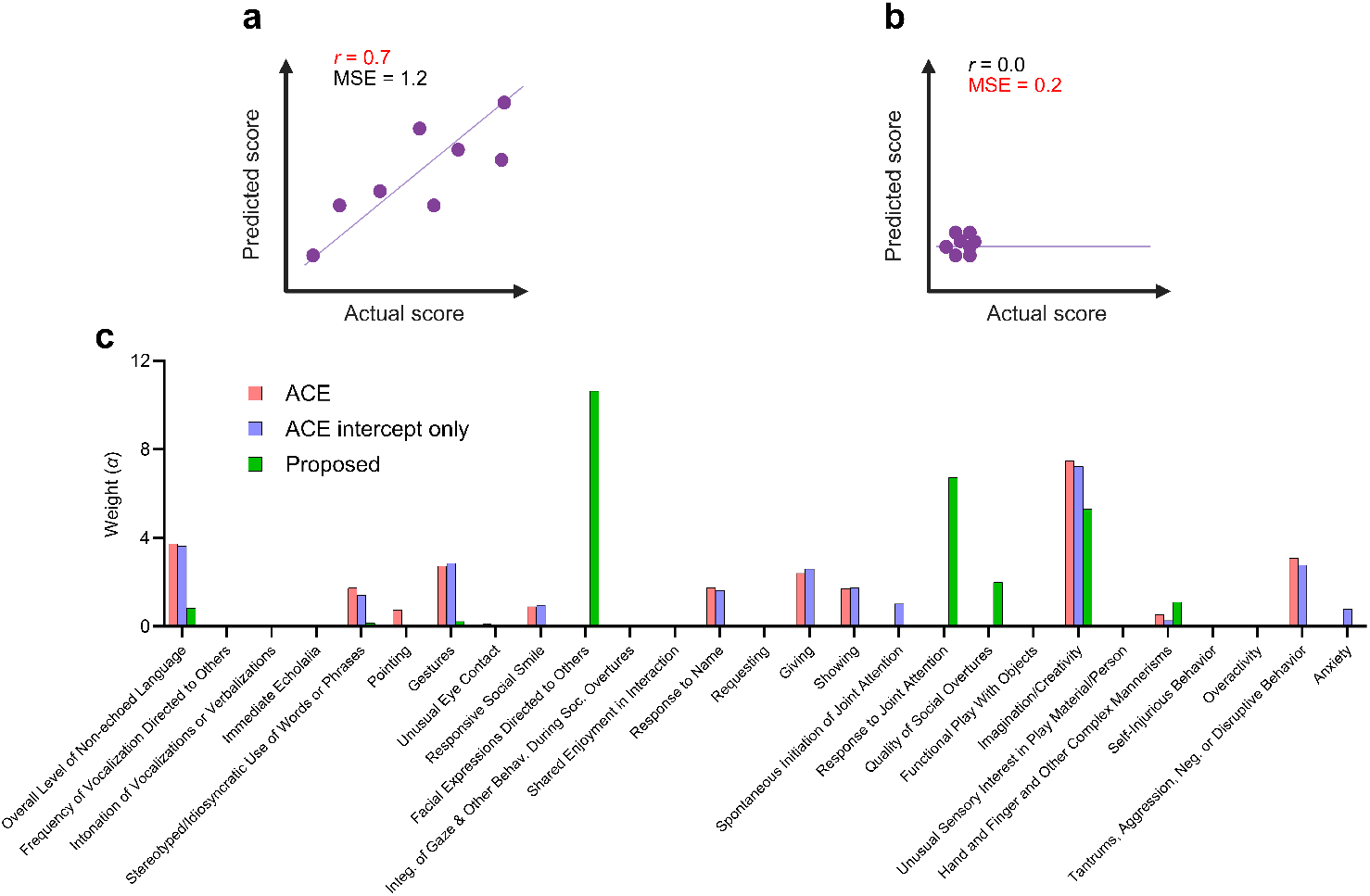
Correlation versus MSE. (a) Maximizing the Pearson correlation coefficient *r* to 0.7 recovers a severity score that distinguishes patients – i.e., separates patients out on the diagonal line – even when the actual values of the score on the *x*-axis vary significantly. However, this strategy leads to a large MSE of 1.2. (b) Minimizing the MSE to 0.2 minimizes error but prefers a stable severity score that only takes on small values on the *x*-axis. The predicted and actual scores are therefore stable but fail to distinguish patients (*r* = 0). (c) We predicted ASD severity at 2 years of age in a group of genetically high-risk patients at 6 months of age by minimizing the MSE with the ACE algorithm. ACE recovered the weights in red that are nearly identical to the weights of the intercept-only model in blue. Maximizing the correlation recovers a very different weights in green.

We provide a concrete application in Figure 2 (c), where we try to predict ASD severity at 2 years of age in a group of genetically high-risk patients at 6 months old; we will elaborate on all approaches and datasets later, but we still seek to motivate the issue here. The prediction problem is known to be *extremely* challenging because social and language capabilities change dramatically during infancy. The ACE algorithm (with a non-negative constraint) minimizes the MSE but yields the weights in red that are nearly equivalent to the intercept-only model in blue; ACE thus simply recovers a minimally variable score for the high-risk group as a whole, rather than distinguish between patients. In contrast, maximizing the correlation with the proposed method using the same data leads to a very different severity score in green than the intercept-only model because correlation emphasizes distinguishing patients over minimizing absolute error.

## 4. Sparse Canonical Outcome Regression

### 4.1. First score

We now describe the proposed Sparse Canonical Outcome REgression (SCORE) algorithm that non-negatively weights the items in a clinical rating scale, where larger values denote greater severity, to generate a summary severity score that differentiates between patients in a high-risk group across time. We thus want to maximize the correlation between a set of *p* baseline predictors *X*_0_ and a set of *q* outcome measures *Y* (*t*_*i*_) across *m* time points *t*_*i*_ ∈ {*t*_1_, …, *t*_*m*_}. We assume missingness at random (MAR) throughout, where the missingness indicators, one for each variable in *Y* (*t*_*i*_), are independent of *Y* (*t*_*i*_) for all *t*_*i*_ given baseline predictors (Rubin, 1976). We also use bold upper case letters like **Y** (*t*_*i*_) to denote a matrix of samples of *Y* (*t*_*i*_), **X**_0_ to denote a matrix of samples of *X*_0_, and **X**_0_ (*t*_*i*_) to denote the rows of **X**_0_ indexed by the non-missing sample indices of **Y** (*t*_*i*_). We require **Y** (*t*_*i*_) and **X**_0_ (*t*_*i*_) to be column-centered at each time point.

SCORE extracts *k* ≤ *p* summary scores. The *j*^th^ score has weights *α* _*j*_ for the rating scale items, and weights *β* _*j*_ = {*β* _*j*_ *t*_1_, …, *β* _*j*_ (*t*_*m*_)} for the predictors, where each *β* _*j*_ (*t*_*i*_) is a column vector for score *j* at time point (*t*_*i*_). We approximate the following population-level objective for the first score (*j* = 1):

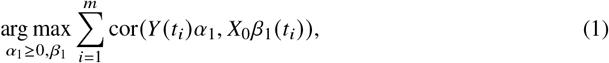

where the non-negativity constraints *α*_1_ ≥ 0 enhance clinical interpretability by naturally favoring a sparse solution (Slawski and Hein, 2013) and ensuring that *Y* (*t*_*i*_) *α*_1_ remains a severity score. The correlation maximization is reminiscent of canonical correlation analysis (CCA), but we maximize a sum of correlations across time rather than a single correlation at a specific time point. Further, the correlations depend on a set of parameters *α*_1_ that remain fixed across time, while *β*_1_ (*t*_*i*_) can differ between time points. We chose this formulation because we want to find a single set of weights *α*_1_ to generate a maximally predictable summary score *Y* (*t*_*i*_) *α*_1_ across time; a dynamic summary score like *Y* (*t*_*i*_) *α*_1_ (*t*_*i*_) would need to be recomputed across time points and may therefore prove too cumbersome for resource-constrained mental health systems or require computational support for clinical use.

We can rewrite Expression (1) using samples as:

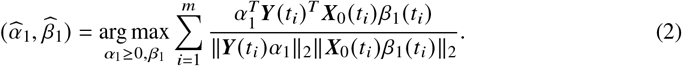

This is non-convex due to the dependence of the denominator on *α*_1_ and *β*_1_. However, we can optimize it using **blockwise coordinate ascent**, alternating between optimizing *α*_1_ and *β*_1_, which guarantees monotonic improvement and convergence to a stationary point under mild conditions (Tseng, 2001). We provide pseudocode in Algorithm 1.

We first fix *α*_1_ and optimize for *β*_1_. As a result, ∥**Y** (*t*_*i*_) *α*_1_∥ _2_ in the denominator of Equation (2) is constant, so we can equivalently solve:

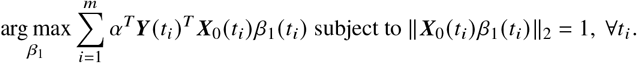

#### Algorithm 1

Sparse Canonical Outcome REgression (SCORE)

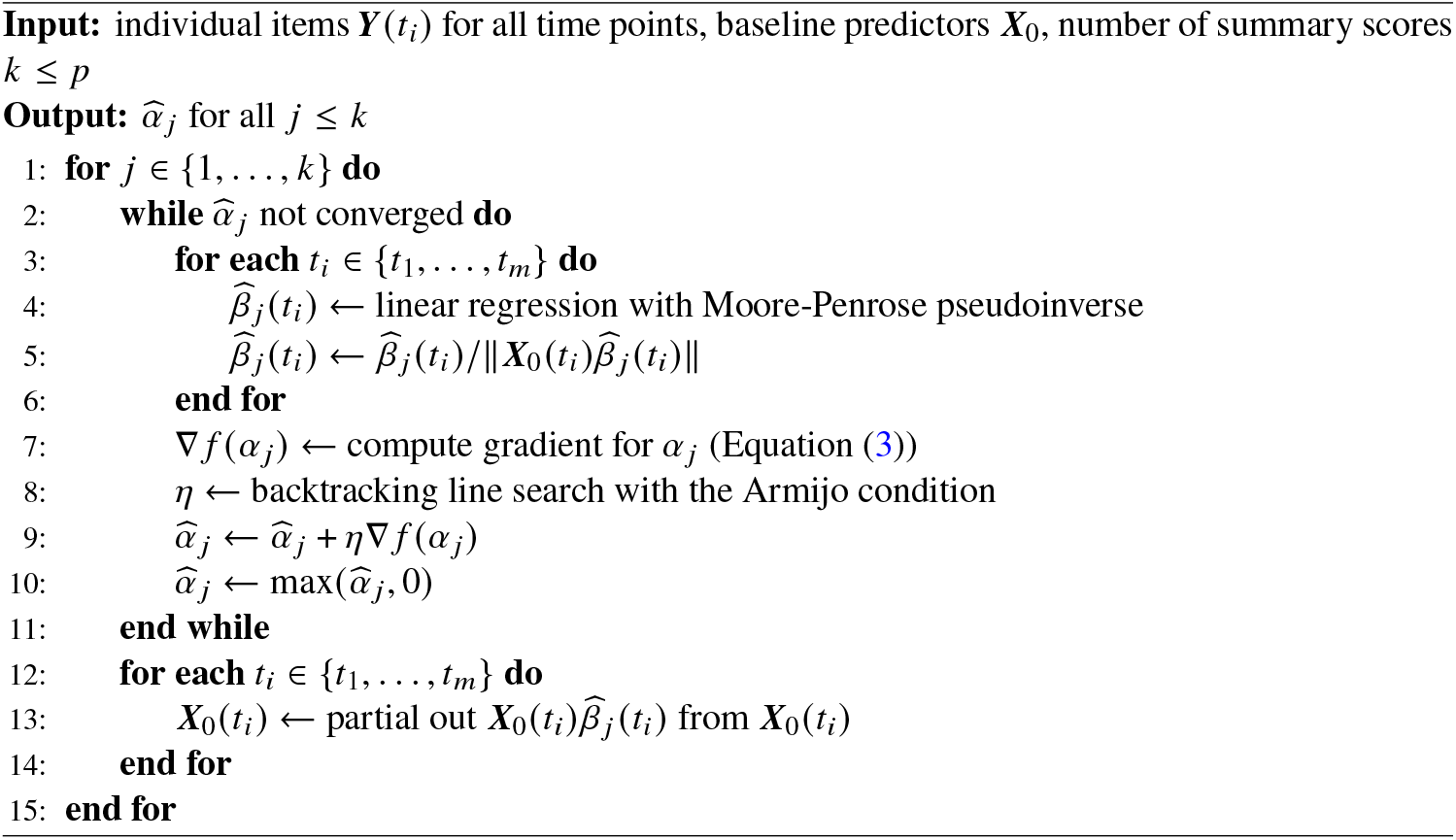

The coefficients *β*_1_ (*t*_*i*_) can differ at each time point, so we instead optimize the following at each time point:

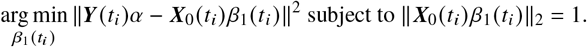

This is a constrained linear regression, which we solve by regressing **Y** (*t*_*i*_) *α*_1_ on **X**_0_ (*t*_*i*_) *β*_1_ (*t*_*i*_) for each time point in Line 4 of Algorithm 1, and then projecting the solution back onto the feasible set to satisfy the constraint ∥**X**_0_ (*t*_*i*_) *β*_1_ (*t*_*i*_) ∥ _2_ = 1 in Line 5. If the linear regression admits multiple solutions, then we prefer coefficients *β*_1_ (*t*_*i*_) with the minimal *L*_2_ norm via the Moore-Penrose pseudoinverse (Golub and Van Loan, 2013).

We next fix *β*_1_ and solve for *α*_1_, or equivalently optimize:

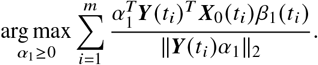

We cannot solve this analytically like with *β*_1_ because *α*_1_ remains fixed across time points. We therefore instead use gradient ascent with projection. Let *ν*_*i*_ = **X**_0_ (*t*_*i*_) *β*_1_ (*t*_*i*_). The gradient of the above expression corresponds to:

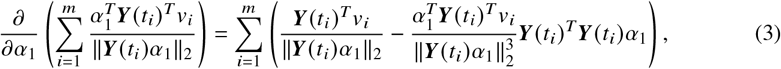

which we compute in Line 7. We then update *α*_1_ in Line 9 after determining the optimal step size *η* by a backtracking line search with the Armijo condition (Armijo, 1966) in Line 8. Finally, we project *α*_1_ back onto the feasible set in Line 10. We iterate through the optimization of *α*_1_ and *β*_1_ until *α*_1_ converges to a stationary point. Let 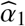 correspond to the final weights of the first summary score, and likewise 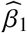 to the final coefficients of the predictors.

### 4.2. Subsequent scores

The first summary score 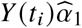 is the most predictable score from baseline according to the correlation coefficient. However, we wish to extract all predictable summary scores as well. CCA traditionally handles this scenario by enforcing uncorrelatedness between 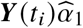 and subsequent combinations of the outcomes 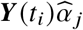 for all 1. CCA also enforces uncorrelatedness between 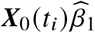 and subsequent combinations of the predictors 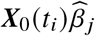 for all *j* > 1. However, we cannot always enforce uncorrelatedness between 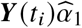 and 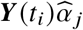 if we wish to keep 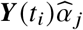 a strict severity score where 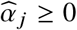.

Strict uncorrelatedness may also not always align with the needs of high-risk patients in clinical management. Imposing the constraint – even approximately – could exclude meaningful symptom patterns that overlap with the first score but still offer distinct predictive value that clinicians might find actionable. For instance, if 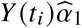 captures social withdrawal in patients at high risk for psychosis, a subsequent score 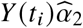 reflecting intolerability to stress might share some correlation with the first score but remain critical for identifying patients needing targeted relaxation techniques. Forcing uncorrelatedness thus risks discarding such overlapping but clinically relevant signals.

We instead seek to identify all summary scores that are predictable from *X*_0_, regardless of their degree of uncorrelatedness, and leave their utility open to expert judgment. This ensures that the algorithm captures the full spectrum of predictable scores. To achieve this, we first solve Expression (2) and then partial out 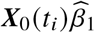 from **X**_0_(*t*_*i*_) in Line 13 by linearly regressing each column of **X**_0_(*t*_*i*_) on 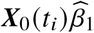 and then taking the residuals. This process removes the linear influence of 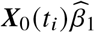 from **X**_0_(*t*_*i*_) by projecting onto the space perpendicular to 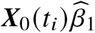. As a result, we can now isolate the maximal sample correlation between **X**_0_(*t*_*i*_) and **Y** (*t*_*i*_) independent of 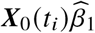 by solving Expression (2) with the new set of baseline predictors. This process yields 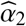 and 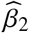 and thus the second summary score 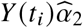. Repeating the above process *k* ≤ *p* times ultimately creates a set of *k* summary scores 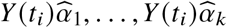 for all time points. Clinicians can then assess each score’s utility based on its predictive strength and clinical context, rather than based on a rigid statistical criterion like uncorrelatedness. This approach thus respects the heterogeneity of clinical management by empowering clinicians to intervene when prediction is reliable and clinically important. The entire SCORE algorithm ultimately runs in *O* (*kmnp*^2 +^ *km p*^3^ + *kmnq*^2^), where *n* denotes the maximal sample size per time point (Appendix Section 7.1).

### 4.3 Predicting a specific time point

Longitudinal data with more than two time points enables additional types of analyses. We do not always wish to recover severity scores that are maximally predictable across a time period from baseline data. Instead, we may also want to identify severity scores that are maximally predictable at a specific time point using all previous time points (Figure 3). For example, a formal diagnosis of ASD usually waits until a patient reaches at least 2 years of age, when language and social skills become more reliable (Lord et al., 2012). However, we may be able to predict *some* symptoms of ASD at 2 years of age during infancy, so we wish to identify summary scores predictable specifically at 2 years of age. We modify the SCORE algorithm to recover severity scores that are maximally predictable at a specific time point in Appendix 7.2.

**Figure 3.**
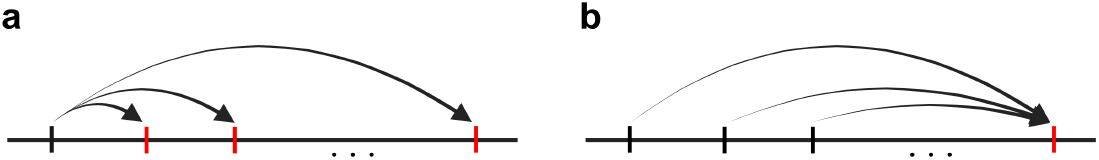
SCORE can handle two related problems. (a) We most commonly want to learn summary scores that are prognostic across all time points in a given time-period (red ticks) using baseline data (black tick). (b) However, there are clinical situations where we want to learn summary scores that are predictable at a specific time point (red tick) using any past time point (black ticks).

## 5. Results

### 5.1. Comparator Algorithms and Clinical Scores

SCORE is the only algorithm that learns maximally correlated severity scores across longitudinal time points. However, we can compare SCORE to other algorithms that learn non-negatively weighted combinations of outcome variables:

1. **Non-negative Canonical Correlation Analysis (NCCA)** (Sigg et al., 2007; Mackey, 2008) is the most similar method to SCORE and thus regarded as the main competitor. NCCA initially finds a non-negative weighting of the outcome variables and a (positive or negative) linear transformation of the predictor variables that maximizes their correlation. NCCA does not account for longitudinal data, so we run it separately for each time point. As a result, the algorithm is similar to SCORE for the first canonical variates, but it does not account for shared information across time points. NCCA then diverges farther from SCORE by imposing a deflation scheme that approximately enforces uncorrelatedness between subsequent canonical variates; in contrast, SCORE iteratively computes residuals only among the predictors.
2. **Alternating Conditional Expectation (ACE)** (Friedman, 1991) finds a non-negative weighting of the outcome variables and a linear transformation of the predictor variables that minimize the MSE, while keeping the variance of the transformed outcomes constant. ACE does not directly maximize the correlation except when *q* = 1. The algorithm alternates between regressing the outcome items on the predictors, and then regressing the predictors on the items. We run the algorithm separately for each time point.
3. **Non-negative Matrix Factorization (NMF)** (Lee and Seung, 2000) finds a low-rank approximation of the outcome variables **Y** (*t*_*i*_) ≈ **W**^*T*^ **H**, where both **W** and **H** have non-negative entries. We obtain a severity score by performing a rank one factorization and then combining the items in **Y** (*t*_*i*_) using the non-negative weights in **H**. We run the algorithm separately for each time point. Note that NMF is an unsupervised method and therefore gives the same severity score regardless of **X**_0_(*t*_*i*_).

We also compare SCORE against the **total score and all clinical subscores** of the clinical rating scales. We compute the total score by summing the scores of each individual item, and the clinical subscores by summing the scores of prespecified items that summarize distinct aspects of the measured condition. For example, the Autism Diagnostic Observation Schedule Version 2 (ADOS-2) Toddler Module can be subdivided into scores related to communication, social interaction, play, restricted/repetitive behaviors, and behavioral regulation (Lord et al., 2012).

### 5.2 Evaluation

Recall from Section 3 that we are clinically interested in maximizing the **correlation across all time points** between learned summary scores and the predictors, because most regression metrics like the MSE are biased towards selecting stable symptoms that yield precise predictions but do not differentiate between high-risk patients. We therefore evaluate the algorithms using Pearson’s correlation coefficient *r* between 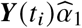 and 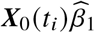 for the first severity score for each time point. We then compute the correlation between 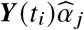 and 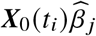 for the other summary scores, after setting **X**_0_ (*t*_*i*_) to its residuals for SCORE (Line 13), or to the generalized deflation of **X**_0_ (*t*_*i*_) for NCCA. Note that NCCA outputs multiple optimal transforms of the outcomes, but ACE only outputs one optimal summary score. As a result, we give ACE the residuals of **X**_0_ (*t*_*i*_) from SCORE for each summary score beyond the first, and then rerun the algorithm with these residuals. Finally, for NMF, we compute the correlation of the original predictors and all of the residuals to the summary score recovered by the algorithm.

We also investigated the minimality of the SCORE algorithm, the **sparsity of the summary scores**, and the **runtime**. We evaluate minimality with **ablation studies** and sparsity using the *L*_0_ norm of the coefficients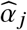 for each time point.

We initialized 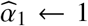 to uniform weights for all algorithms to prevent initialization bias and to ensure that the denominator of Equation (3) remains non-zero for stable optimization. We then use warm starts 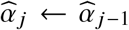 for subsequent scores. None of the algorithms used additional hyperparameters, including ablated variants of SCORE. We did not impute any data.

We statistically compared the performance of each algorithm according to the correlation coefficient, *L*_0_ norm and run-time by drawing 5000 bootstrap samples for each dataset and then running each algorithm on each bootstrapped draw. We assess the algorithms on the approximately 36.8% of samples not drawn in each bootstrap draw. We then compute 95% confidence intervals and compare methods by performing two-sided paired t-tests. All statements made below hold at a Bonferroni corrected threshold of 0.05 divided by the total number of comparator algorithms plus the total number of clinical scores.

### 5.3. Predicting Predictable Psychotic Symptoms

We downloaded data from the North American Prodrome Longitudinal Study Version 3 (NAPLS-3) from the National Institute of Mental Health (NIMH) Data Archive (Dataset ID # 2275) with a limited access data use certificate awarded to author Eric V. Strobl (Addington et al., 2022). This study was a multi-site research initiative focused on understanding the predictors and mechanisms of psychosis development in youth at clinical high risk (CHR). CHR status was determined as individuals with attenuated positive symptoms, brief intermittent psychotic episodes, or genetic risk plus functional decline, indicating elevated psychosis risk without meeting full diagnostic criteria. NAPLS-3 then employed a longitudinal design, conducting clinical and biomarker assessments every two months for the first eight months, followed by clinical evaluations at 12, 18, and 24 months. During the SIPS interview, a clinician rated each patient using the Scale Of Prodromal Symptoms (SOPS), a 19-item rating scale used to assess the severity of subthreshold symptoms of psychosis across four domains: positive (P), negative (N), disorganization (D) and general (G) (Miller et al., 1999). We list each item of the SOPS on the *x*-axis of Figure 4 (a).

**Figure 4.**
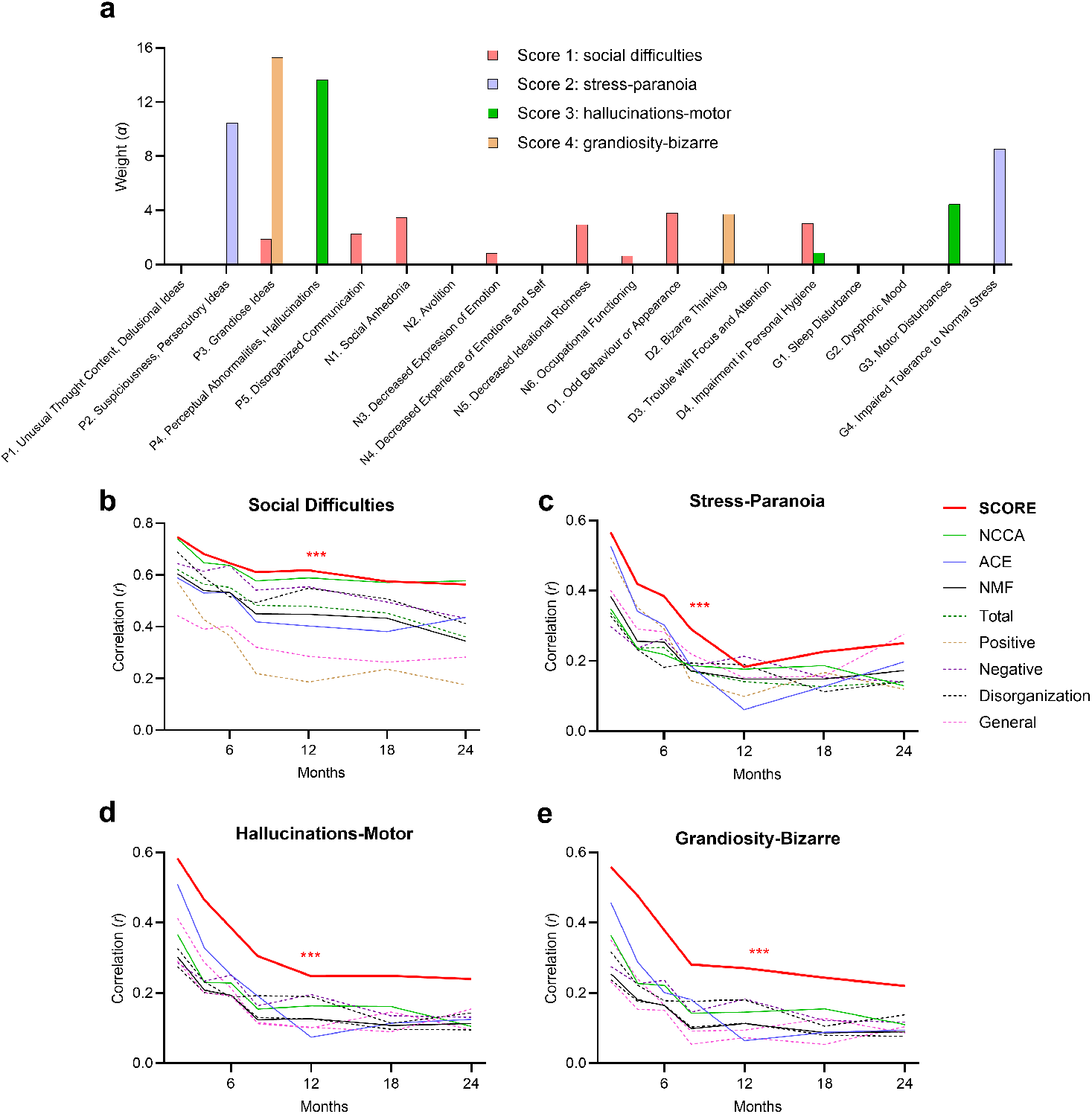
Results of predicting psychotic symptoms over a two-year period. (a) The weights of the top 4 severity scores discovered by SCORE are sparse. (b) SCORE outperformed all other algorithms in the first recovered score across all time points, except NCCA at 24 months. (c,d,e) Similarly, SCORE outperformed all other algorithms in the second, third, and fourth scores across all time points. Note that subfigures (b)-(e) share the same legend, and the SCORE algorithm has a 95% confidence error band of the mean, although it is not visible in each subfigure due to the large number of bootstrap samples. * *p* < 0.05, ** *p* < 0.005, *** *p* < 0.0005 from nearest competitor.

The clinical task is to differentiate CHR patients whose psychotic symptoms will worsen from those whose symptoms will not over a two-year time period using age, biological sex, and the individual SOPS items at baseline.

We thus analyzed data from CHR patients who had complete data at baseline for age, biological sex and all items of the SOPS, as well as complete data at at least one of the 7 follow-up time points for all items of the SOPS. We list key characteristics of this cohort at baseline in Appendix Table 1.

#### 5.3.1. Analysis of Outputs

We plot the first severity score recovered by SCORE in red in Figure 4 (a). The score combined multiple items in the SOPS related to social difficulties. For example, the score had large weights on disorganized communication, social anhedonia, odd behavior or appearance, decreased ideational richness, and impairment in personal hygiene. This social difficulties score cut across traditional symptom domains by combining items across positive, negative, and disorganization symptoms. Moreover, the score achieved the largest correlation coefficient with baseline predictors across all follow-up time points (Figure 4 (b)). NCCA came in at a close second but fell short particularly at 4, 8 and 12 months because it did not use the full set of longitudinal samples across all time points like SCORE. We conclude that SCORE learned the most predictable severity score from baseline data.

We likewise plot the second, third, and fourth severity scores recovered by SCORE in Figure 4 (a). These scores only depended on 2-3 items each and were thus much sparser than the first score with little overlap; the sparsity results with bootstrapped samples were similar (Appendix Figure 6 (a)). The sparse severity scores also achieved the highest correlation coefficients with predictors across nearly all time points compared to all other methods (Figure 4 (c)-(e)). The margin between SCORE and the other algorithms widened with increasing numbers of score extractions; SCORE completely dominated all comparators by the third and fourth severity scores because existing methods were either not designed to extract all predictable severity scores (NMF, ACE, total score, subscores), or impose uncorrelatedness constraints that limit predictability (NCCA). We conclude that SCORE outperformed existing methods by a larger margin with increasing numbers of extractions. We discuss the clinical implications of these results in Section 6.

Ablation studies revealed that SCORE requires both the correlation objective and the non-negativity constraints to achieve maximal performance (Appendix Figure 8). Finally, SCORE had a shorter run-time than NCCA and completed within 3 seconds in this dataset (Appendix Figure 7 (a)).

### 5.4. Predicting Predictable ASD Symptoms

We next pushed the algorithms to their limits by assessing their ability to predict ASD from infancy, a notoriously challenging task where noise overwhelmingly overshadows signal due to the variable course of social and language development in infants. Algorithms that are not carefully designed tend to break in this regime by either capturing only group-level patterns (e.g., ACE in Section 3), or by relying on all rating scale items and thus compromising clinical interpretability. We downloaded data from the Infant Brain Imaging Study (IBIS) study in the NIMH Data Archive (Dataset ID # 19) (Hazlett et al., 2017). IBIS is a multisite research initiative that primarily recruits infants who have an older sibling diagnosed with ASD, since these genetically high-risk siblings have approximately a 20% chance of developing ASD themselves. IBIS seeks to identify early brain and behavioral markers that predict a diagnosis of ASD to enable earlier interventions that improve patient outcomes.

Investigators in the IBIS study administered the Autism Observation Scale for Infants (AOSI) at 6 and 12 months of age. The AOSI consists of semi-structured, play-based observations over 18 different tasks related to social interaction, communication, and sensory-motor functioning (Bryson et al., 2008). Each task is rated on a 4-point Likert scale. The AOSI emphasizes early risk indicators rather than a definitive diagnosis of ASD.

Investigators in IBIS then administered the ADOS-2 Toddler Module at 24 months of age. This module focuses on observable social, communicative, and play-based behaviors (Lord et al., 2012). The IBIS dataset contains the ratings of 21 items from the ADOS-2 Toddler Module at 24 months of age; we list the individual items along the *x*-axis of Figure 5 (a). In contrast to the AOSI, the ADOS-2 Toddler Module assesses symptoms of ASD that can support a clinical *diagnosis*. As a result:

**Figure 5.**
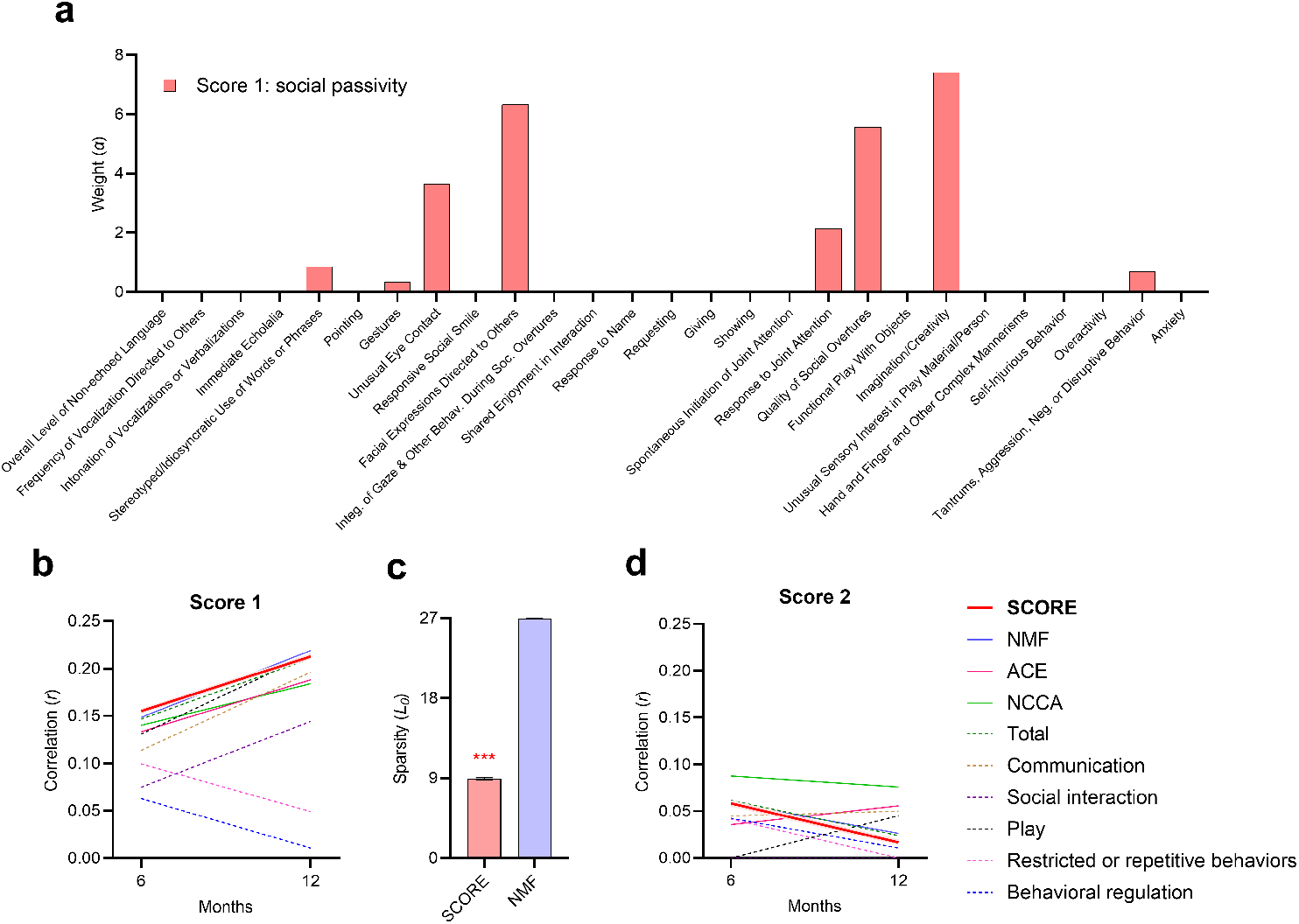
Results for predicting symptoms of ASD at 2 years of age. (a) SCORE discovered a sparse weighting of the items of the ADOS-2 Toddler Module involving symptoms related to decreased social overtures and imaginary play, which we collectively call social passivity. (b) SCORE and NMF both outperformed all other methods as averaged over both time points. (c) SCORE recovered a three times more sparser solution than NMF for the first summary score. (d) No method detected a significantly predictable score (*r* > 0.1) beyond the first score.

The clinical task is to differentiate genetically high risk patients who will exhibit symptoms of ASD from those who will not at 2 years of age using exact age, biological sex and the individual AOSI items at approximately 6 months and 12 months of age.

Notice that we are predicting symptoms at 2 years of age, rather than across a time period, as discussed in Section 4.3.

#### 5.4.1. Analysis of Outputs

We ran the algorithms with exact age, biological sex and all items of AOSI at 6 and 12 months in order to predict the ADOS-2 Toddler Module items at 24 months. We provide detailed baseline cohort characteristics at 6 and 12 months in Appendix Table 2.

The SCORE algorithm extracted one predictable severity score with the largest weights on quality of social overtures, imagination/creativity, unusual eye contact and facial expressions directed towards others (Figure 5 (a)). Imagination and creativity facilitate social overtures in toddlerhood through pretend play and shared activities (Smits-van der Nat et al., 2024). Impairments in the above items thus hinder toddlers from initiating and sustaining social interactions. We therefore collectively summarize the deficits in the above items as social passivity, where the child fails to pursue shared experiences with others.

SCORE achieved the largest correlation coefficient at month 6 for the first summary score (Figure 5 (b)). Across both time points, SCORE surpassed all comparators except NMF, with which it performed comparably (*t* = 0.23, *p* = 0.82). Moreover, SCORE recovered weights that were approximately three times sparser than the weights recovered by NMF for the first summary score (Figure 5 (c)). NMF unfortunately recovered dense solutions in general across all summary scores despite its non-negativity constraint (Appendix Figure 6 (b)). Moreover, the weights learned by NMF were nearly uniform and difficult to clinically interpret (Appendix Figure 10), in contrast to the targeted sparse weights of SCORE. We conclude that SCORE learned the sparsest, clinically interpretable summary score while achieving the highest predictability across time in the IBIS dataset. Ablation studies further confirmed the necessity of the components of SCORE in the IBIS dataset (Appendix Figure 9). SCORE had a shorter run-time than NCCA and usually completed within one second (Appendix Figure 7 (b)).

None of the algorithms discovered scores beyond the first with a correlation coefficient above 0.10 (Figure 5 (d)). This outcome likely stems from the inherent challenge of predicting ASD symptoms using the AOSI. Unlike other domains like imaging, where outcomes often exhibit a priori predictability, forecasting ASD may lack inherent reliability and render additional predictable patterns beyond social passivity elusive.

## 6. Discussion

We aimed to identify all clinical severity scores that distinguish the future trajectories of high-risk patients. As a result, we developed a carefully designed algorithm called Sparse Canonical Outcome REgression (SCORE) that achieves this objective by learning severity scores that maximize predictability based on the correlation coefficient. Maximizing the correlation enables the scores to differentiate between patients in the high-risk category. Furthermore, iterating over the residuals of the predictors allows the algorithm to discover multiple severity scores. SCORE thus robustly learns multiple predictive models while also ensuring maximal predictability of the outcomes, succeeding even in extremely challenging tasks like ASD prediction.

The severity scores identified by SCORE diverge significantly from metrics commonly employed in the clinical literature. For example, investigators determine psychosis conversion using SIPS, where patients must attain a severity score of 6 on at least one of the five positive symptoms of SOPS (P1-P5 in Figure 4 (a)) for at least one hour per day, four days per week, across one month (Miller et al., 2003). Investigators may waive frequency requirements if positive symptoms trigger severely disorganized or dangerous behaviors. The SIPS criteria thus prioritize positive symptoms. In contrast, SCORE uncovered more predictable severity scores related to social difficulties, stress-paranoia, motor symptoms, and grandiosity that extend beyond a near-exclusive focus on positive symptoms.

The severity scores generated by SCORE provide a compelling basis for clinical intervention. For instance, many patients in the psychosis prodromal phase display unusual social behaviors often dismissed as a typical adolescent phase (Ballon et al., 2007). Predicting the persistence of these symptoms can justify early social skills training, even if patients do not meet psychosis conversion criteria (Kelsven et al., 2022). Likewise, anticipating elevated stress-paranoia or grandiosity can support the use of cognitive behavioral therapy (CBT) techniques to alleviate paranoia, anxiety, and distorted thinking (Hagen et al., 2013). SCORE also revealed that AOSI scores from infancy predict challenges with social overtures, imagination, and facial expressions by age two. Although these correlations were weaker, they support parent training approaches to enhance positive social behaviors and communication (Landa, 2018). Thus, SCORE enables researchers to pinpoint predictable scores that justify milder clinical actions emphasizing prevention rather than the treatment of fully developed mental illness. As a result, the scores may also require less predictive certainty compared to interventions targeting diagnosable conditions.

Note that SCORE explicitly generates *static, interpretable* severity scores that can be precomputed and easily memorized for clinical use, unlike traditional machine learning models that often demand real-time deployment. The severity scores thus eliminate the need for ongoing algorithmic computation in clinical settings. Clinicians can also readily incorporate the scores into routine assessments by focusing on symptoms tied to the positive weights of predictable scores – such as social difficulties in psychosis risk or passivity in ASD risk – without requiring specialized tools. The proposed approach therefore imposes no technical burden, making SCORE well-suited for resource-constrained mental health systems.

SCORE exhibits certain limitations despite its strengths. The algorithm currently identifies linear combinations of predictors and outcomes to ensure straightforward clinical interpretability. Substituting a non-linear method, such as kernel ridge regression, into SCORE may increase its predictive accuracy. Additionally, SCORE generates multiple severity scores by iteratively analyzing the residuals of the predictors, but a method that detects all predictable severity scores without altering the predictor set could preserve predictive strength across scores. This approach, nevertheless, demands a new definition of “distinctiveness” for severity scores. Third, SCORE deliberately learns static severity scores that remain maximally predictable over time, which suits gradually evolving psychiatric illnesses. However, developing severity scores that adapt over time may prove beneficial for rapidly changing conditions, such as those in intensive care units. Future research should thus prioritize integrating non-linearity, reducing predictor transformations, and devising time-varying severity scores.

In conclusion, we developed the SCORE algorithm to predict and distinguish the future trajectories of high-risk patients. This algorithm excels in addressing complex mental illnesses that a single total severity score cannot adequately capture, instead offering detailed characterization through multiple individual items. In practice, SCORE tends to uncover severity scores tied to subtle symptomatologies, potentially justifying early clinical action even with mild interventions.

## Data Availability

All data produced are available online at https://nda.nih.gov/.

### 7. Appendix

#### 7.1. Computational Complexity

We now derive the computational complexity of the SCORE algorithm. The linear regression in Line 4 takes *O* (*np*^2^ + *p*^3^) time, which we repeat *m* times. The gradient step in Line 7 is dominated by the multiplication **Y** (*t*_*i*_) ^*T*^**Y** (*t*_*i*_) which takes *O* (*nq*^2^) time for *m* time steps. Suppose the algorithm requires at most *L* iterations of the while loop in Line 2 for 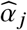 to converge. Then, the convergence takes *O* (*L* (*mnp*^2^ +*m p*^3^)) + *O* (*Lmnq*^2^) time corresponding to the linear regression and the gradient step, respectively. Next, each regression in Line 13 takes *O* (*n*) time, repeated for *p* variables in **X**_0_ and *m* time points; thus, computing the residuals takes *O* (*mnp*) time in total. Note that we repeat all of the above steps at most *k* times. As a result, the total computational complexity of SCORE is *O* (*k L* (*mnp*^2^ + *m p*^3^)) + *O* (*k Lmnq*^2^) + *O* (*kmnp*). If we treat *L* as a constant, then we arrive at the final complexity *O* (*kmnp*^2^ + *km* + *p*^3^ *kmnq*^2^). The modified SCORE algorithm below has the same complexity.

#### 7.2. Modified SCORE

We can modify the SCORE algorithm to recover severity scores that are maximally predictable at a specific time point. We obtain the *j* ^th^ severity score by optimizing:

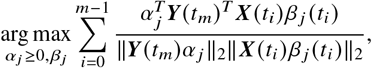

where we have replaced **Y** (*t*_*i*_) with the rating scale items only at the last time point **Y** (*t*_*m*_), and we have replaced the predictors at baseline **X**_0_ (*t*_*i*_) with predictors **X** (*t*_*i*_) collected at specific time points preceding *t*_*m*_ in time. The gradient then becomes:

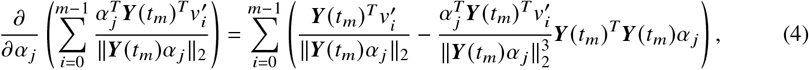

where we now have *ν*_*i*_^′^ = **X** (*t*_*i*_) *β* _*j*_ (*t*_*i*_). We list the modified pseudocode in Algorithm 2, where changes are highlighted.

##### Algorithm 2

Modified SCORE for predicting one time point

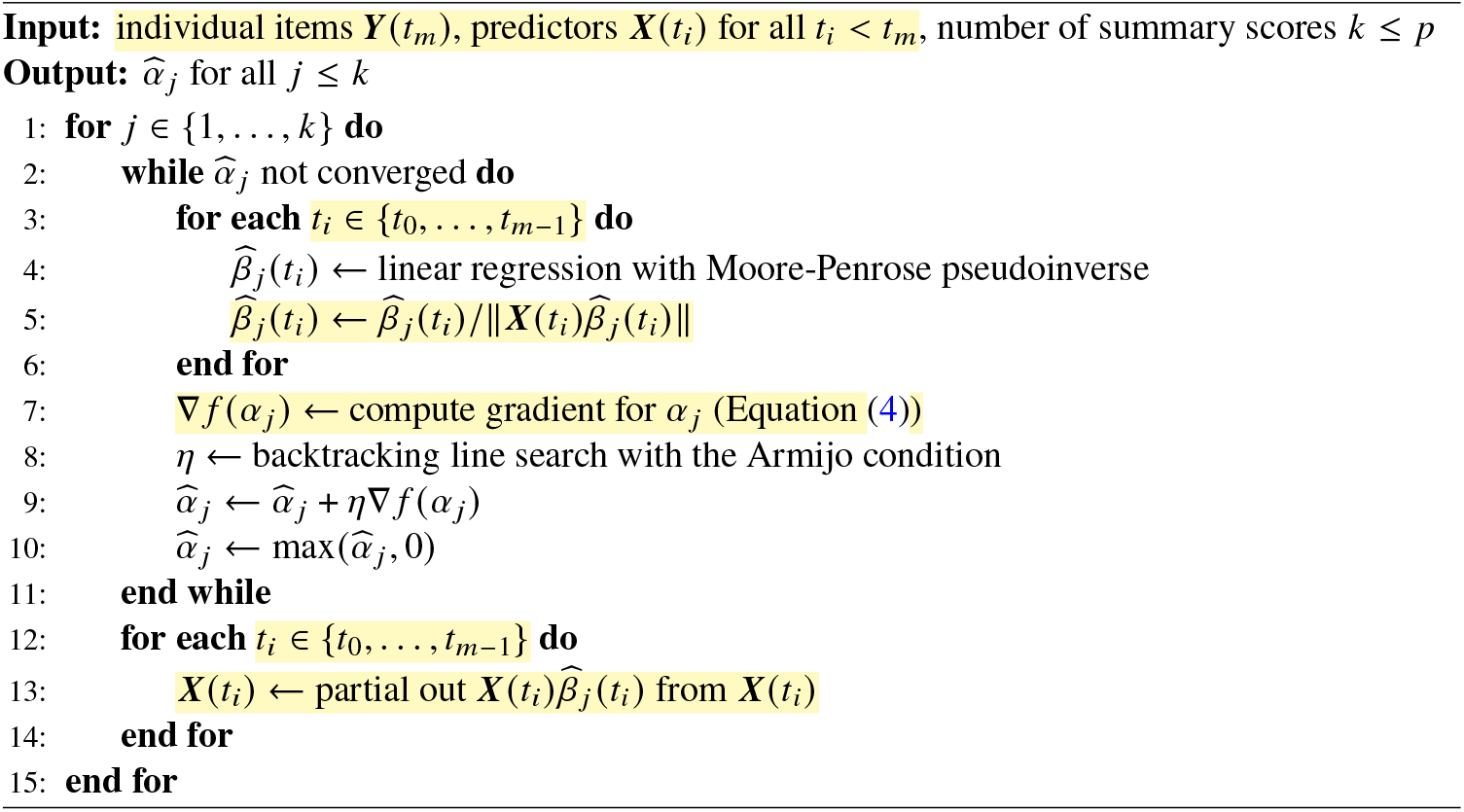

#### 7.3. Cohort Tables

**Table 1:** Characteristics of the NAPLS-3 cohort at baseline, summarized as mean (standard deviation). Patients must have complete data for age, biological sex, and all individual SOPS items at baseline and at at least one follow-up time point.

|  | Baseline |
| --- | --- |
| Number of patients | 292 |
| Age (years) | 19.13 (4.11) |
| Biological sex (male = 1) | 0.52 |
| SOPS items |  |
| P1. Unusual Thought Content, Delusional Ideas | 3.75 (1.09) |
| P2. Suspiciousness, Persecutory Ideas | 3.16 (1.37) |
| P3. Grandiose Ideas | 0.84 (1.30) |
| P4. Perceptual Abnormalities, Hallucinations | 3.16 (1.40) |
| P5. Disorganized Communication | 1.93 (1.49) |
| N1. Social Anhedonia | 2.42 (1.72) |
| N2. Avolition | 2.55 (1.55) |
| N3. Decreased Expression of Emotion | 1.40 (1.41) |
| N4. Decreased Experience of Emotions and Self | 1.87 (1.60) |
| N5. Decreased Ideational Richness | 1.19 (1.28) |
| N6. Occupational Functioning | 2.60 (1.95) |
| D1. Odd Behaviour or Appearance | 0.80 (1.20) |
| D2. Bizarre Thinking | 0.95 (1.19) |
| D3. Trouble with Focus and Attention | 2.55 (1.33) |
| D4. Impairment in Personal Hygiene | 0.86 (1.25) |
| G1. Sleep Disturbance | 2.38 (1.42) |
| G2. Dysphoric Mood | 3.22 (1.62) |
| G3. Motor Disturbances | 0.85 (1.02) |
| G4. Impaired Tolerance to Normal Stress | 2.84 (1.91) |

**Table 2:** Characteristics of the IBIS cohort at the two baseline time points. Patients must have complete data for age, biological sex, and all individual AOSI items at 2 years of age and at at least one baseline time point.

|  | 6 months | 12 months |
| --- | --- | --- |
| Number of patients | 320 | 308 |
| Age (months) | 6.62 (0.76) | 12.56 (0.77) |
| Biological sex (male = 1) | 0.63 | 0.63 |
| AOSI items |  |  |
| Visual tracking | 0.36 (0.71) | 0.46 (0.89) |
| Disengagement of attention | 0.83 (1.15) | 0.21 (0.81) |
| Orients to name | 1.08 (1.23) | 0.62 (1.05) |
| Differential response to facial emotion | 1.79 (2.37) | 1.34 (2.28) |
| Anticipatory response | 0.89 (1.29) | 0.34 (0.81) |
| Imitation of actions | 2.12 (1.94) | 0.51 (1.13) |
| Social babbling | 1.88 (1.07) | 1.10 (1.19) |
| Eye contact | 0.45 (1.00) | 0.27 (0.68) |
| Reciprocal social smile | 0.61 (0.94) | 0.43 (0.72) |
| Coordination of eye gaze and action | 0.16 (0.64) | 0.01 (0.11) |
| Reactivity | 0.35 (0.63) | 0.20 (0.50) |
| Social interest and shared affect | 0.51 (0.82) | 0.31 (0.51) |
| Transitions | 0.13 (0.70) | 0.18 (0.44) |
| Motor control and behavior | 0.28 (0.67) | 0.05 (0.22) |
| Atypical motor behaviors | 0.13 (0.63) | 0.16 (0.68) |
| Atypical sensory behaviors | 0.16 (0.66) | 0.25 (0.87) |
| Engagement of attention | 0.28 (1.12) | 0.19 (1.04) |
| Insistence on having or playing | 0.24 (1.12) | 0.27 (1.06) |
| Social referencing | 1.12 (1.31) | 0.46 (1.13) |

#### 7.4. Sparsity Results

**Figure 6.**
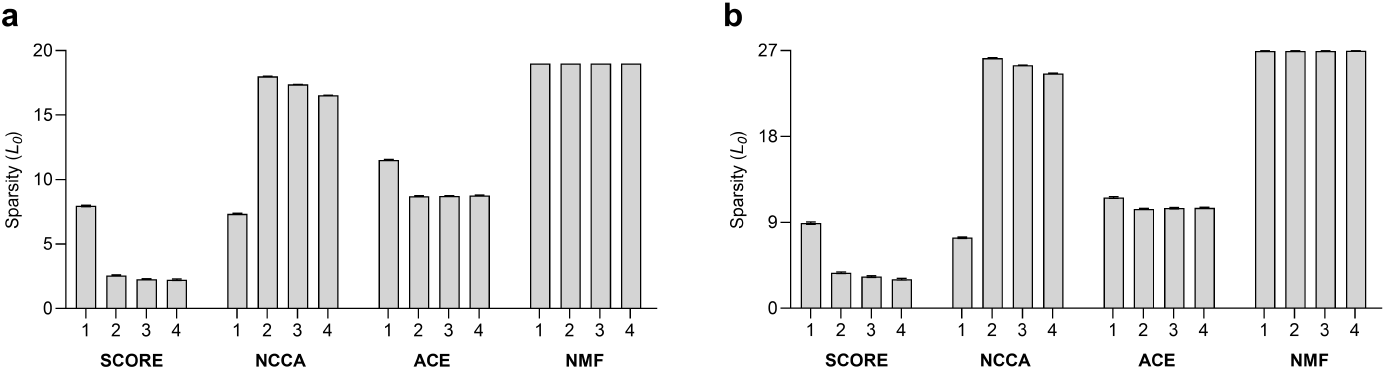
Bar graphs with 95% confidence intervals depicting 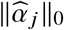 with *j* ∈ {1, 2, 3, 4} averaged across time points for (a) NAPLS-3 and (b) IBIS. Note that SCORE and NMF output the same non-negative weights for all time points, but NCCA and ACE output potentially different non-negative weights for each time point. SCORE and NCCA achieve similar sparsity at the first severity score. SCORE attains the sparsest solution for the second, third and fourth severity scores.

#### 7.5. Timing Results

**Figure 7.**
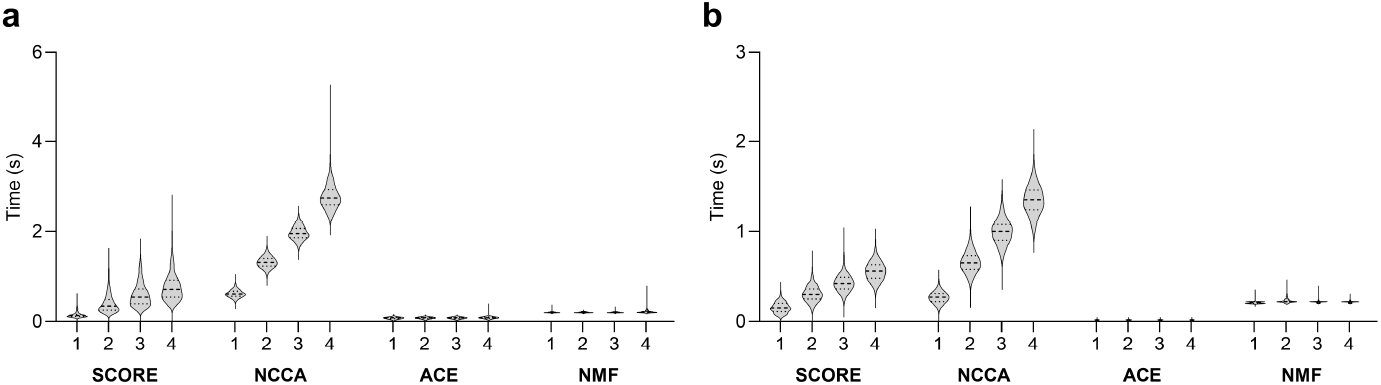
Violin plots summarizing timing results in (a) NAPLS-3 and (b) IBIS for learning 1-4 scores. SCORE took longer than ACE and NMF but required less time than NCCA. All algorithms completed within a few seconds. Lines represent the first, second and third quartiles.

#### 7.6. Ablation Results

We hypothesize that the correlation objective and the non-negativity constraint are both necessary components of the SCORE algorithm. We tested this hypothesis by first removing the sample correlation objective and replacing it with an MSE:

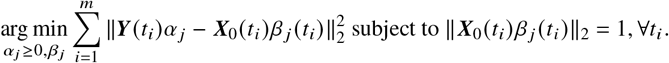

We solve this problem by block coordinate descent, where we (1) fix *α* _*j*_ and solve for *β* _*j*_ as in SCORE, (2) normalize *β* _*j*_ as in Line 5 to prevent convergence to a trivial *α* _*j*_ = *β* _*j*_ = 0 solution, (3) fix *β* _*j*_ and solve for *α*_*j*_ ≥ 0 by running non-negative least squares in place of the gradient update in Lines 7 to 9, and then (4) repeat until convergence.

We also consider a second ablated variant of SCORE, where we remove the non-negativity constraint:

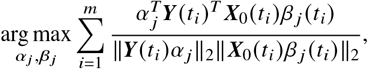

operationalized by removing the projection step in Line 10. We then enforce non-negativity only after we have fully optimized 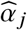 for each *j*. Finally, we consider a combined variant, where we remove both the correlation objective and the non-negativity constraint by optimizing:

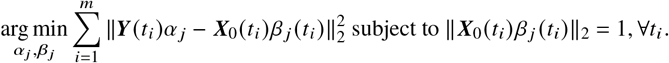

We solve this problem also by block coordinate descent but fix *β* _*j*_ and solve for *α* _*j*_ by running simple least squares.

We plot the results of SCORE and all three variants with the NAPLS-3 dataset in Figure 8. We also plot the IBIS dataset results in Figure 9. We find that removing the correlation objective, non-negativity constraint or both decreases the accuracy of SCORE as assessed by the Pearson correlation coefficient. We therefore conclude that both of these components of SCORE are necessary for the algorithm to achieve maximal performance.

**Figure 8.**
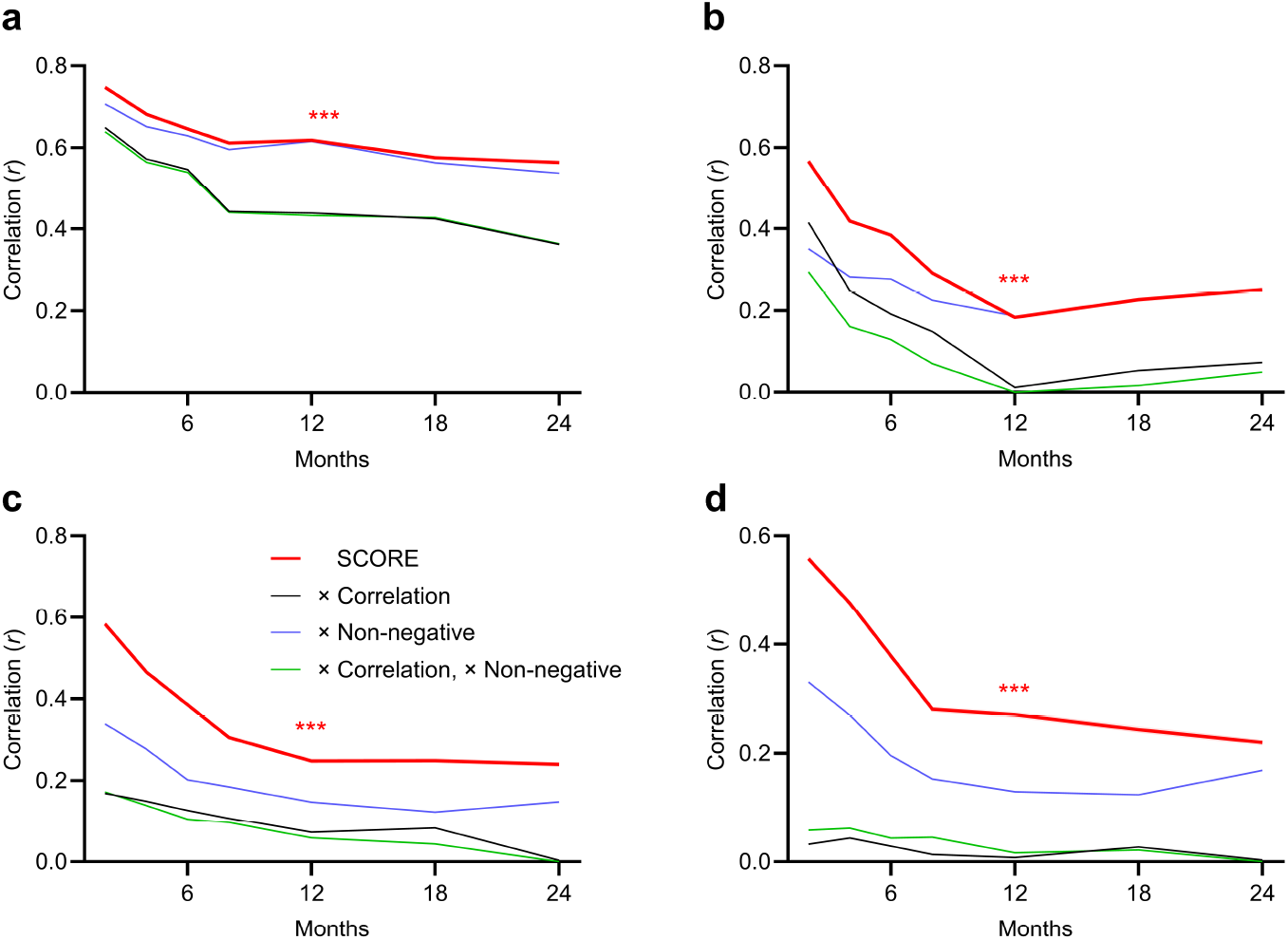
NAPLS-3 ablation results with the correlation objective removed (and replaced by the MSE), the non-negativity constraint removed, and both the correlation objective and the non-negativity constraint removed. Subfigures (a)-(d) correspond to the first through fourth scores recovered by the algorithms. SCORE outperformed all three ablated variants in the NAPLS-3 dataset, indicating that both the correlation objective and the non-negativity constraint are necessary for SCORE to maximize the Pearson correlation on held-out data.

**Figure 9.**
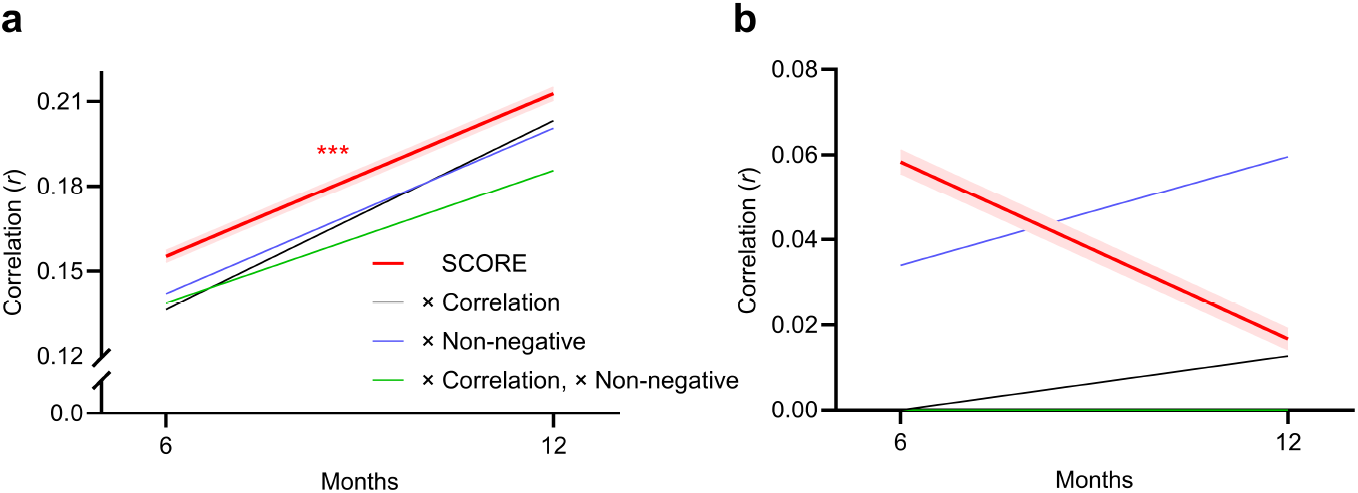
IBIS ablation results. (a) SCORE outperformed all ablated variants with the first score. (b) All methods again achieved low Pearson correlation coefficients (*r* < 0.1) from the second severity score onward.

#### 7.7. Clinical Interpretability in IBIS

**Figure 10.**
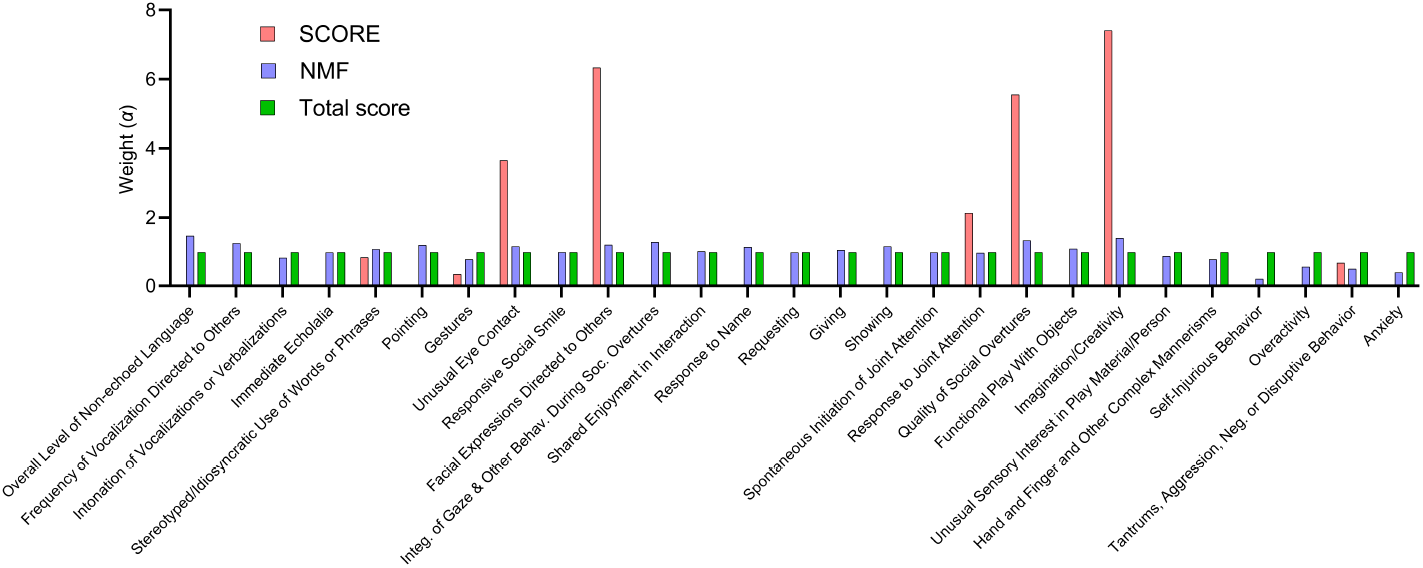
Weights of the first severity score recovered by NMF and SCORE on IBIS. NMF yielded nearly uniform weights that simply mimic the total severity score. As a result, clinicians cannot easily determine the symptoms of ASD predictable from infancy using NMF. In contrast, SCORE produced sparse, peaked weights that highlight specific symptom patterns.

